# COVID-19 Pandemic-related differences in the prevalence of genetic conditions and healthcare utilization and access for children with genetic conditions the United States

**DOI:** 10.1101/2025.04.14.25325831

**Authors:** Abhinav Thakral, Divya Moorthy, Tanya Tandon, Gianluca diUbaldi, Gregg LaFleur, Kusum Viswanathan, Fernanda Kupferman

**Author notes:** Corresponding author: Dr. Abhinav Thakral.

## Abstract

**Purpose:** to assess the pandemic-related differences in the prevalence of diagnosed genetic conditions (GC) in children (0-17 years) and their healthcare access and utilization in the US

**Methodology:** Using the National Survey of Children’s Health (NSCH) data, we compared pre-pandemic (2016-2019) and pandemic (2020-2022) prevalence estimates for diagnosed GC, healthcare utilization (HCU), and unmet health needs (UHN). The prevalence ratios (PR) of HCU and UHN during versus pre-pandemic for different groups were compared using interaction models, adjusted for age, sex, socioeconomic status, and race-ethnicity.

**Results:** Pre-pandemic, GC prevalence was 3.55% rising to 4.51% during the pandemic [PR = 1.27 (1.18, 1.37), p < 0.001]. Pre-pandemic, children with GC had higher HCU [PR = 1.148 (1.129, 1.167), p < 0.001] and UHN [PR = 2.701 (2.225, 3.278), p < 0.001] as compared to children without GC. For children with GC, there was a decrease in HCU [PR = 0.954 (0.931, 0.978), p < 0.001], however, UHN was similar [PR = 1.112 (0.882, 1.402), p = 0.37] during versus pre-pandemic.

**Conclusion:** The prevalence of diagnosed GC increased and disparities in UHN for children with GC persisted during the pandemic.

**What is known about this topic?:** While children with genetic conditions have greater utilization of healthcare services, they also have greater unmet health needs as compared to those without. On top of this disparity, the pandemic disrupted health services.

**What this paper adds?:** The prevalence of diagnosed genetic conditions increased during the pandemic, and while healthcare utilization decreased, unmet health needs remained the same for children with genetic conditions.

## Introduction

Individual genetic conditions are rare, however, taken together, they constitute a substantial minority presenting with a unique aspect of healthcare that is often overlooked or neglected. It is estimated that in 2016-2017, around 2.8 million children aged 0 to 17 years were living with a reported genetic condition including cystic fibrosis, Down syndrome, blood disorders, and other genetic or inherited conditions ^1^. These estimates are likely to be under-representative of the true prevalence of genetic conditions given the barriers to access to genetic testing which limit the number of children with recognized genetic conditions. Such barriers include a lack of awareness on the part of patients and their healthcare providers, a lack of knowledge of genetic services ^2^, a lack of access to genetic service providers and labs, and socioeconomic and racial disparities, amongst others ^3,4^. Similar factors ^5–8^ also pose barriers to healthcare access for children with genetic conditions. In addition, children with genetic conditions often have unique healthcare needs, such as mobility issues, and immunodeficiencies predisposing them to adverse health outcomes during in-person visits in crowded waiting rooms, a requirement for specialized care, amongst others. Unsurprisingly, despite greater requirements for healthcare ^1,9–11^ children with genetic conditions have greater unmet healthcare needs as compared to children without genetic conditions ^1^.

The COVID-19 pandemic was a time of great upheaval and posed several challenges to healthcare access and delivery for all children ^12–16^. These challenges added further to the pre-existing barriers facing children seeking a genetic diagnosis. Furthermore, the pandemic impacted even universal newborn screening programs ^17–19^, a common avenue for genetic diagnosis. Thus, the pandemic may have significantly prolonged the time to diagnosis for children with genetic conditions and potentially reduced the prevalence of children with a recognized genetic condition. However, the initial decline in healthcare utilization during the pandemic was followed by a recovery period^20^. Furthermore, telemedicine use is reported to have increased during the pandemic ^21,22^. Due to the complex interplay of pandemic-related factors, it is difficult to assess the impact of the pandemic, if any, on delays in genetic diagnosis or the prevalence of children with recognized genetic conditions. To our knowledge, this impact has not yet been investigated.

In addition to adding delays for children seeking genetic diagnosis, the pandemic also impacted children with diagnosed genetic conditions. For instance, studies on individual genetic conditions have highlighted worsening mental health and food insecurity suffered by patients living with such conditions ^23–26^. In terms of healthcare access and utilization, there is limited data for children with diagnosed genetic conditions. Though Macaluso et al demonstrated that the pandemic negatively affected access to care for people with rare conditions, the data are limited for children under 18 years, who were underrepresented in the study ^27^.

Thus, we conducted the present study to assess the pandemic-related differences in the prevalence of children with diagnosed genetic conditions and the healthcare access and utilization for children with genetic conditions in the United States. We hypothesized that there were pre-existing disparities in healthcare access and unmet health needs for children with genetic conditions and that the pandemic exacerbated these disparities.

The performance of the healthcare system in times of crises can be gauged by how the system treats its vulnerable and minoritized individuals. By providing national-level estimates on the impact of the pandemic on this medically vulnerable section of society, the study will provide a metric to inform health institutions and policymakers.

## Methods

### Study Design

Cross-sectional

### Dataset

To study the COVID-19 pandemic-related differences in the prevalence of genetic conditions, we used the National Survey of Children’s Health (NSCH) data from 2016-2022. The NSCH methodology is published by the Census Bureau ^28^. Briefly, the Census Bureau conducts the survey annually between July of that year and January of the following year. The survey sample is a stratified random sample covering the 50 states and the District of Columbia. Participants are contacted through web, email, mail, and telephone. Only one child is selected from each household.

Each child within the survey is given a survey weight to obtain stable national and state-level estimates. These sampling weights are adjusted for several factors, including demographics, non-response, as well as for households with more than one child.

The response rates for the survey were between 39-43% for the years 2016-2022. The response rate during the pandemic years was similar to that before the pandemic ^29^. Missingness for genetic conditions in the dataset was around 1% and was not associated with the pandemic time-period (supplementary table 1). Therefore, we excluded the missing values from the analysis. Due to the low missingness, the magnitude of selection bias resulting from missing values should be low.

The sampling strategy for NSCH changed in 2021-2022 and the harmonized datasets were released recently.

### Variables

#### Primary variables

Our primary variables of interest were “genetic condition”, “healthcare utilization”, and “unmet healthcare needs”.

Regarding Genetic conditions, we computed the variable based on responses to four survey questions. The survey had questions related asking “Has a doctor or a health care provider ever told you that your child has …” We used the responses to the questions for three medical conditions namely cystic fibrosis, Down syndrome, and other genetic or inherited conditions. We computed the variable “genetic condition” as “Yes” if the response to any question was “Yes”.

For healthcare utilization, we used the response to the question “In the past 12 months, did you visit any healthcare provider for this child?”

For unmet healthcare needs, we used the response to the question “In the past 12 months, did you not receive healthcare for your child that you felt that the child needed”. People who responded “yes” to this question were further asked about what type of healthcare was not received. We used the responses to this question for the type of healthcare not received.

#### Time-periods and multi-year weighting

For the pandemic period, we included the years 2020-2022 in the pandemic period and divided the weights for the pandemic period by 3, per the analytic guidelines from NSCH.

For the pre-pandemic period, we combined the years 2016-2019 and divided the weights by 4, per the same analytic guidelines by NSCH.

#### Confounders

Our potential confounders were age, sex, race-ethnicity, and socioeconomic status (SES). Race/ethnicity was based on self-identified race and Hispanic ethnicity. If the respondent on the survey indicated “Hispanic ethnicity”, the race/ethnicity was given the value “Hispanic (any race)” otherwise, the value of the race/ethnicity was the self-identified race of the respondent. As there were fewer than 30 samples for certain years for American Indian and Alaska Native (AIAN) children, Hawaiian or Other Pacific Islander children, and children with unknown/more than one race, the estimates were unstable for these groups. Hence, children from these groups were included in one group for stable estimates.

We classified children into socioeconomic status (SES) based on family income. We used the Poverty Income Ratio (PIR), the ratio of the family income to the Federal Poverty Line, to classify children into 4 categories, with children with PIR<1.3 classified as low, PIR>=1.3, and <= 1.85 as lower middle, >1.85 and <=3.5 as middle, and > 3.5 as upper SES. These classifications were based on the eligibility for federal SNAP (Supplemental Nutrition Assistance Program), eligibility for the WIC program (Women, Infants, and Children), and CDC analytic guidelines (22).

#### Statistical analysis

We determined the unadjusted weighted prevalence of genetic conditions. Next, we determined the weighted estimates of prevalence of healthcare utilization and unmet health needs, including which needs were unmet, for each of the two time periods for children with and without genetic conditions. To compare the ratio of prevalence of genetic conditions during versus before the pandemic, we used log-binomial regression. For the ratio of prevalence of healthcare utilization and unmet health needs during versus before the pandemic, we separately calculated the ratios for children with and without genetic conditions using log-binomial regression.

For comparing pandemic-related differences in healthcare utilization and unmet healthcare needs for children with and without genetic conditions, we used two interaction models with “healthcare utilization”, and “unmet health needs” each regressed on a genetic condition, pandemic time-period and the interaction term between the two, adjusting the analysis for age, sex, race-ethnicity, and socio-economic status as potential confounders. We conducted an “analysis of variance” of these interaction models to assess if the pandemic-related differences in healthcare utilization, and unmet health needs were different for children with and without genetic conditions. We attempted log-binomial regression for these models, however, as the model failed to converge, we used Poisson regression with robust standard errors. We presented the results as ratios of pandemic and pre-pandemic prevalences.

All analyses were conducted using survey weights, strata, and primary sampling units to reflect national-level estimates for non-institutionalized children in the United States, per the NSCH analytic guidelines ^30^. All statistical tests were done using the Survey R package ^31^.

The analyses were considered exempt by the institutional review board of Brookdale Hospital.

## Results

Table 1 shows the baseline population characteristics of the survey population before and during the COVID-19 pandemic. The median age slightly decreased from 10.0 to 9.0 years. The sex distribution remained stable, with males constituting around 52% and females about 48% in both periods. There were significant shifts in socioeconomic status, with a slight decline in the proportion of individuals from high socioeconomic backgrounds and a rise in those from low and lower-middle groups. Racial and ethnic composition also changed, with increases in the proportions of African American, Asian, Hispanic, and other racial groups, while the percentage of White participants declined from 70.2% to 65.4%. The unweighted prevalence of genetic conditions in the survey sample rose modestly from 3.9% to 4.5%.

**Table 1:** Survey population characteristics before and during the pandemic.

| Population Characteristics | levels | Pre-pandemic<br>N (%) | During Pandemic<br>N (%) | P value* |
| --- | --- | --- | --- | --- |
| Age in years | Median (IQR) | 10.0 (5.0 to 14.0) | 9.0 (4.0 to 14.0) | <0.001* |
| Sex | Male | 68100 (51.7) | 76483 (51.8) | 0.683** |
|  | Female | 63674 (48.3) | 71289 (48.2) |  |
| Socioeconomic Status | High (PIR >3.5) | 64309 (48.8) | 70687 (47.8) | <0.001** |
|  | Low (PIR < 1.3) | 21037 (16.0) | 25524 (17.3) |  |
|  | Lower Middle (PIR 1.3-1.85) | 11989 (9.1) | 13529 (9.2) |  |
|  | Middle (PIR 1.85-3.5) | 34439 (26.1) | 38032 (25.7) |  |
| Race-ethnicity | African American | 8209 (6.2) | 9510 (6.4) | <0.001** |
|  | Asian | 6741 (5.1) | 8545 (5.8) |  |
|  | Hispanic | 15138 (11.5) | 21087 (14.3) |  |
|  | Others | 9179 (7.0) | 12055 (8.2) |  |
|  | White | 92507 (70.2) | 96575 (65.4) |  |
| Genetic Condition | Yes | 5074 (3.9) | 6611 (4.5) | <0.001** |

\*P value is from the Kruskal-Wallis test, \*\*P-value is from Chi-squared test.

The weighted prevalence of diagnosed genetic conditions was 3.55% (3.36, 3.75) during the pre-pandemic time-period and 4.51% (4.3, 4.73) during the pandemic. The weighted prevalence ratio of diagnosed genetic conditions during the pandemic versus before the pandemic was 1.27 (1.18, 1.37), p < 0.001.

Table 2 compares the weighted healthcare utilization and unmet health needs among children with and without genetic conditions before and during the COVID-19 pandemic. Across both periods, children with genetic conditions had higher healthcare utilization rates and greater unmet needs than those without. Pre-pandemic, 93.2% of children with genetic conditions had a healthcare visit compared to 82.6% without; this decreased slightly during the pandemic to 89.5% and 80.8%, respectively. Unmet healthcare needs were consistently higher among children with genetic conditions—rising from 8.6% pre-pandemic to 9.2% during the pandemic—while children without genetic conditions saw a similar increase from 2.8% to 3.3%.

**Table 2:**
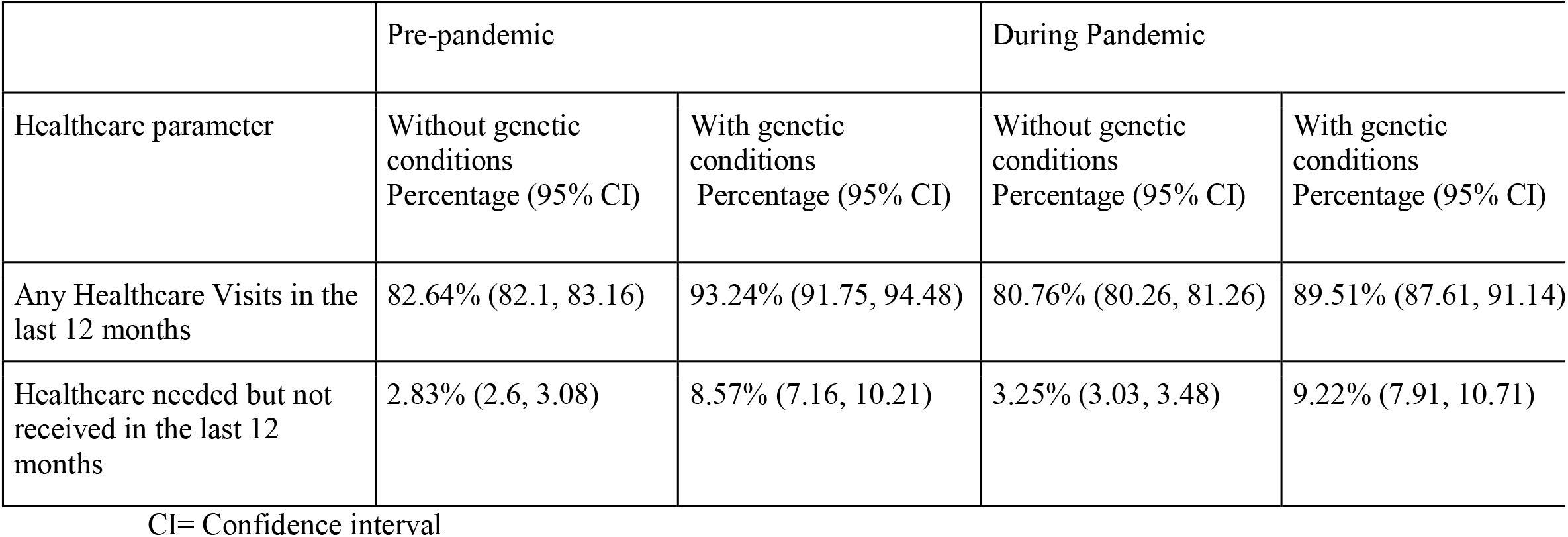
pre-pandemic and pandemic percentage prevalences (with 95% confidence interval) of healthcare utilization, unmet health needs, and specific types of unmet health needs for children with and without genetic conditions.

Table 3 summarizes the results of the interaction models in terms of the pre-pandemic ratio of prevalence of healthcare utilization and unmet health needs for children with and without genetic conditions (first two columns), and the pandemic-related ratios of prevalence for children with genetic conditions (next two columns). While children with genetic conditions had a higher prevalence of having any healthcare visits, they also had higher unmet health needs as compared to children without genetic conditions before the pandemic. During the pandemic, there was a decrease in the prevalence of healthcare visits for children with genetic conditions, however, the unmet health needs did not change for children with genetic conditions compared to before the pandemic.

**Table 3:**
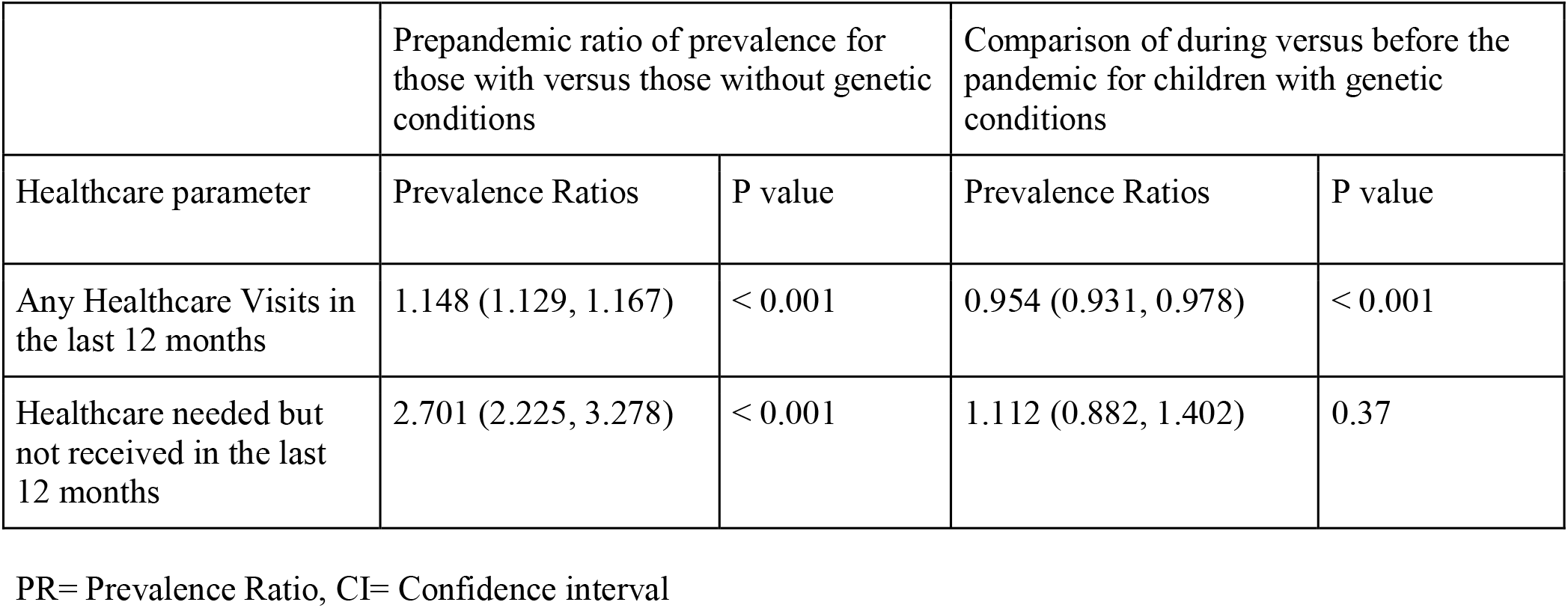
Weighted prevalence ratios (with 95% confidence interval) of healthcare utilization and unmet health needs from the interaction models. The first two columns show the pre-pandemic ratio of prevalence for children with versus without genetic conditions and the P values from the weighted interaction models, while the next two columns show the pandemic versus pre-pandemic ratios of the prevalence for children with genetic conditions and the P values from the weighted interaction models.

|  | Prepandemic ratio of prevalence for those with versus those without genetic conditions |  | Comparison of during versus before the pandemic for children with genetic conditions |  |
| --- | --- | --- | --- | --- |
| Healthcare parameter | Prevalence Ratios | P value | Prevalence Ratios | P value |
| Any Healthcare Visits in the last 12 months | 1.148 (1.129, 1.167) | < 0.001 | 0.954 (0.931, 0.978) | < 0.001 |
| Healthcare needed but not received in the last 12 months | 2.701 (2.225, 3.278) | < 0.001 | 1.112 (0.882, 1.402) | 0.37 |

PR= Prevalence Ratio, CI= Confidence interval

From the analysis of variance of the interaction models, the pandemic-related difference in healthcare utilization (p = 0.097) and unmet health needs (p = 0.738) were similar for children with and without genetic conditions.

## Discussion

Our study on the impact of the pandemic on children with genetic conditions found that there was an increase in the prevalence of diagnosed genetic conditions from 3.55% before the pandemic to 4.51% during the pandemic. Children with genetic conditions were more likely to utilize healthcare services and also have greater unmet health needs. We also found that healthcare utilization decreased, and unmet health needs did not change for children with genetic conditions. The pandemic-related differences in these parameters were comparable between the groups.

This rise in the prevalence of diagnosed genetic conditions during the pandemic could be due to a complex interplay of several competing factors. Firstly, the initial decrease in access to healthcare services during the pandemic was subsequently followed by a recovery with an increase in the use of telemedicine ^20–22^. Telehealth in genetic services has been shown to be non-inferior to in-person genetic visits, and is increasingly being recognized in the field of genetic practise ^32–34^. An increase in access to telehealth could have opened greater slots and improved access to genetic services for previously underserved populations. Secondly, there has been a trend towards an increase in the use of next-generation sequencing technology ^35^, which has been shown to improve the yield of genetic diagnoses for several previously underdiagnosed conditions ^36,37^. Thirdly, some of the rise in prevalence could also be attributed to anecdotal evidence of parents spending more time with their children during the pandemic leading to a greater recognition of symptoms of childhood genetic conditions, which in turn led to increased consultations and testing via telehealth services.

There was a pandemic-related decrease in access to healthcare and an increase in unmet healthcare needs for all children during the pandemic, including those with and without genetic conditions. While our interaction model did not detect a difference between the groups, we did identify pre-existing disparities in unmet health needs, which is similar to a previous single-year analysis by Lichstein et al ^1^. One would assume that an increase in access to telehealth during the pandemic could have helped address these gaps. Unfortunately, our analysis suggests that these disparities persisted during the pandemic. Other researchers have identified worsening mental health and food insecurity among individuals with different genetic conditions ^23–26^. Our findings, taken together with those from other studies highlight that it is pertinent to address this gap in the unmet healthcare needs for children with genetic conditions and will require urgent and multi-pronged action from healthcare providers, policymakers, social workers, and other allies in child healthcare teams. There is an urgent need to develop innovative and integrative models of healthcare to address these disparities.

Notably, we used a different definition of genetic conditions compared to those used previously^1^. While many blood disorders, including sickle cell anemia and hemophilia are inherited, the response to the question related to presence of a blood disorder can introduce a misclassification bias in the findings, hence we excluded the question from our definition, although it has previously been used^1^. We expect our findings to be more specific, although they might exclude the responses for which the parents might be unaware of the genetic basis of their child’s inherited blood condition.

Our study has the strength of being nationally representative with a large sample size. However, our study also has certain limitations. Our study is based on survey data and is hence subject to biases such as non-response bias and recall bias. The non-response bias with NSCH is small and we suspect that this bias should not have overtly influenced our results. Regarding recall bias, though caregiver reports of genetic conditions for their children have been validated ^38,39^, there is still a possibility that differences in caregiver understanding of the genetic etiology of their child’s medical condition may cause either under or over-reporting of genetic diagnosis. Finally, while we investigated the pandemic-related differences in prevalence, healthcare utilization, and unmet health needs for children with genetic conditions, it is not possible to draw a causal interpretation from our findings, as a complex interplay of pandemic and non-pandemic-related factors would have contributed to the differences we found.

To conclude, despite the limitations, our study provides an estimate of the pandemic-related differences in the prevalence of diagnosed genetic conditions and the differences in healthcare utilization, and unmet health needs for such children. We identified an increase in the prevalence of genetic conditions during the pandemic, possibly stemming from a complex interplay of factors such as wider availability of telehealth and wider availability of genomic sequencing technology leading to better detection of genetic conditions. There was a decrease in healthcare utilization for all children though the unmet health needs for children with genetic conditions did not increase during the pandemic, the prepandemic disparities persisted during the pandemic. The unmet health needs of children with genetic conditions represent an urgent call that must be addressed through a concerted effort from healthcare providers, institutions, and policymakers.

## Acknowledgements

none

## Tables and Figures

**Supplementary Table 1:**
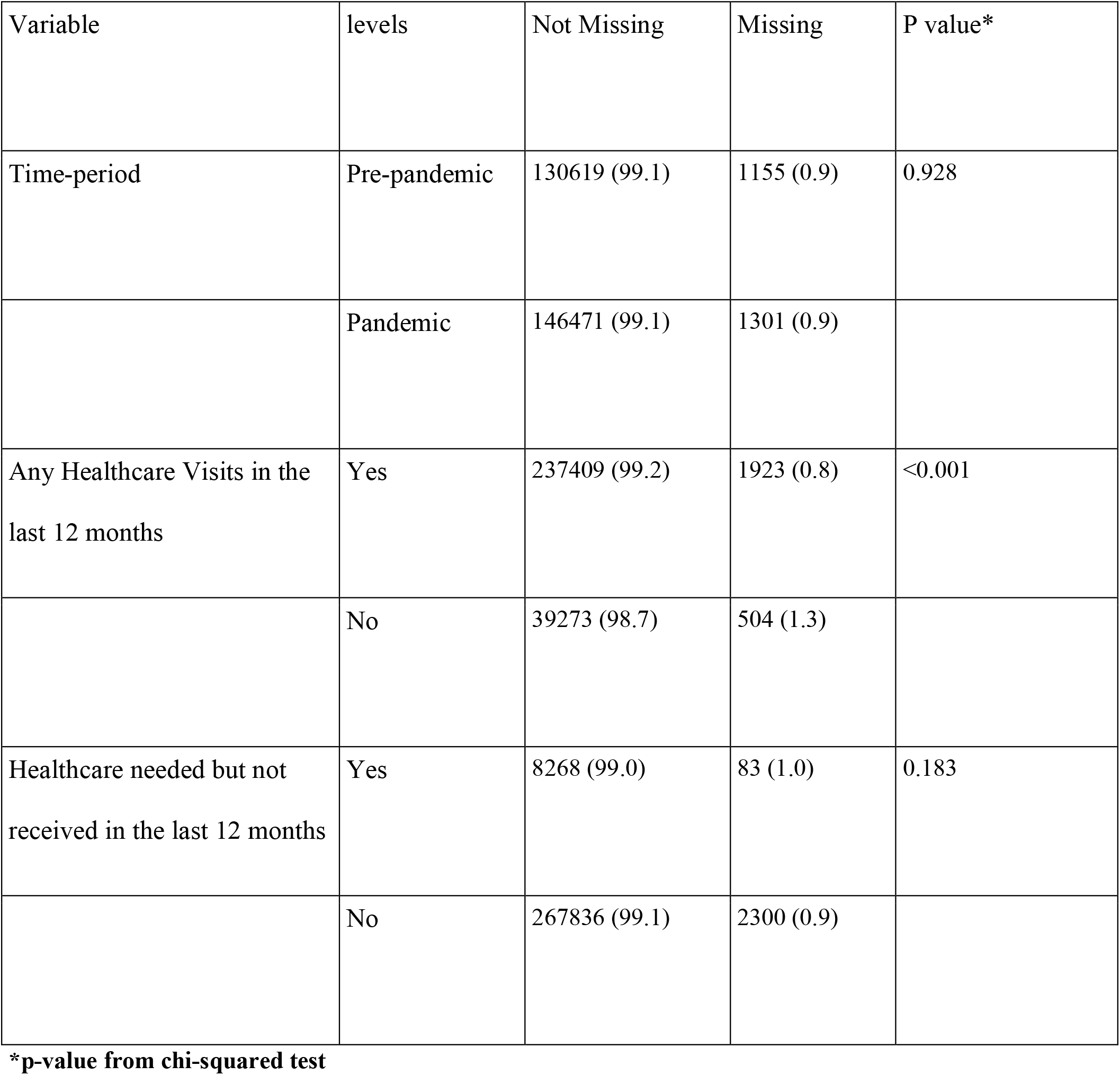
Association of missingness in the “genetic conditions” variable with different variables of interest.

## Data Availability

Data used in the current study is available publicly from the NSCH website (https://www.census.gov/programs-surveys/nsch/data/datasets.html)

## Author Contributions

Conceptualization: AT, DM, TT, FK, KV, Formal analysis: AT, Methodology: AT, DM, TT, Project administration FK, KV, Supervision: FK, KV, Writing-original draft: AT, TT, DM, GD, GL, Writing-review & editing: AT, TT, DM, GD, GL, FK, KV

## Patient Consent

Consent was needed as this is a secondary analysis of survey data

## Ethics Declaration

The project was deemed exempt from the Institutional Review Board at Brookdale Hospital.

